# Psychological outcomes of 12-15-year-olds with gender dysphoria receiving pubertal suppression in the UK: assessing reliable and clinically significant change

**DOI:** 10.1101/2023.05.30.23290763

**Authors:** Susan McPherson, David E.P. Freedman

**Author notes:** Corresponding author: Professor Susan McPherson, School of Health and Social are, University of Essex, Colchester, UK.

## Abstract

The evidence base for psychological benefits of GnRHA for adolescents with gender dysphoria (GD) was deemed “low quality” by the UK National Institute of Health and Care Excellence. Limitations identified include inattention to clinical importance of findings. This secondary analysis of UK clinical study data uses Reliable and Clinically Significant Change approaches to address this gap. The original uncontrolled study collected data within a specialist GD service. Participants were 44 12-15-year-olds with GD. Puberty was suppressed using “triptorelin”; participants were followed-up for 36 months. Secondary analysis used data from parent-report Child Behaviour Checklists and Youth Self-Report forms. Reliable change results: 15-34 percent of participants reliably deteriorated depending on the subscale, time point and parent versus child report. Clinically significant change results: 27-58 percent were in the borderline (subclinical) or clinical range at baseline (depending on subscale and parent or child report). Rates of clinically significant change ranged from 0-35%, decreasing over time towards zero on both self-report and parent-report. The approach offers an established complementary method to analyse individual level change and to examine who might benefit or otherwise from treatment in a field where research designs have been challenged by lack of control groups and low sample sizes.

## Introduction

The UK Gender Identity Development Service (GIDS) was established in 1989 providing therapeutic assessment and psychological intervention for children and young people experiencing gender dysphoria (GD). An early audit of the service found that of the first 124 referrals to the service, the majority had complex social and/or psychological difficulties in addition to gender dysphoria (Di Ceglie, Freedman, McPherson, & Richardson, 2002). At that time, GIDS was based on a psychotherapeutic service model based within an NHS specialist mental health Trust in London. When children reached adulthood (at least 16) and wanted to pursue medical interventions such as puberty blockers, cross-sex hormones or surgery, they would be referred to adult services in a hospital setting.

Puberty blockers (GnRHa treatment) are a form of medical intervention, licenced for use to retard puberty in young people with precocious puberty, that have additionally been prescribed for people experiencing GD, previously restricted to those aged 16 years and over. The intervention had been provided to younger children in the Netherlands since the late 1990s (Cohen-Kettenis & van Goozen, 1998). From around 2009 a number of countries including USA and Australia began to permit its use in this younger age group (Carmichael et al., 2021a). In 2009 the British Society of Endocrinology and Diabetes published a statement advocating their use in a cautious and carefully managed way, monitored within a research context (Barnes, 2023).

Also in 2009, the service commissioning structure at the UK specialist GD service changed from a London based service receiving extra-contractual referrals on a case-by-case basis to a nationally commissioned specialist service (Barnes, 2023). Referrals began to rapidly increase from 2009 to 2019, while levels of complex social and psychological difficulties in the client group remained high (Holt, Skagerberg, & Dunsford, 2016). Controversy concerning referrals for puberty blockers in this service has been ongoing since 2011 when treatment was first offered to 12-15-year-olds taking part in research and subsequently offered routinely from 2014. The main argument for the introduction of puberty blockers in the UK for this age group had been their potential to relieve psychological distress.

> “A key purpose of GnRHa treatment is to pause puberty, to avoid a deterioration in wellbeing and allow for further exploration of a young person’s feelings about their gender identity and their wishes for the future, without the pressure or distress which may come from further unwanted bodily changes” (Gender Identity Development Service, n.d.)

However, in a high-profile case, the UK High Court ruled in 2020 that under 16s could not legally consent to puberty blockers, a decision later overturned by the Court of Appeal, decisions which led to ongoing disruptions and uncertainty for patients and families. In June 2023, NHS England announced it would no longer provide puberty blockers outside of research.

To inform an ongoing NHS review of services in this area, the UK’s National Institute for Health and Care Excellence (NICE) published an evidence review (NICE, 2020) on “Gonadotrophin releasing hormone analogues for children and adolescents with gender dysphoria”. Using GRADE methodology to assess study quality, the majority of evidence was deemed to provide “very low” certainty across a wide range of outcomes including mental health, quality-of-life, bone density and cognitive function. The review followed standard methodologies employed by NICE for identifying evidence, appraising methodological quality and synthesising outcomes. A further literature surveillance report was produced (NHS England, 2023) to determine if the evidence review required updating. The surveillance report found only two additional studies that might “materially affect the conclusions” of the 2020 evidence review.

In relation to mental health and psychosocial outcomes (the focus of the present study), the evidence review summarised outcomes from three studies. Neither of the two studies in the surveillance report were considered likely to materially affect conclusions regarding mental health or psychosocial outcomes. The three studies relevant to mental health were two prospective longitudinal studies (Costa et al., 2015; de Vries, Steensma, Doreleijers, & CohenLJKettenis, 2011) and one cross-sectional study (Staphorsius et al., 2015). One of these was a study of 70 teenagers which found statistically significant reduction in depression; no statistically significant impact on anger or anxiety; statistically significant improvements on internalising and externalising problems measured by the Child Behaviour Checklist (CBCL) and Youth Self Report Form (YSR); and statistically significant improvement in global functioning measured by the Children’s Global Assessment Scale (CGAS) (de Vries et al., 2011). Another found no improvements on CGAS scores among 201 teenagers over 6 months (Costa et al., 2015). The cross-sectional study (Staphorsius et al., 2015) involved 40 adolescents but the NICE review concluded that the analysis of CBCL outcomes was unclear.

The NICE review noted that the studies reviewed were generally “not reliable and changes could be due to confounding, bias or chance” (NICE, 2020). Common issues include lack of comparison or control groups, small samples, poor reporting of physical and mental comorbidities and concomitant treatments, variability and poor reporting of ages children started treatment, unclear analyses and, in particular, little interpretation of clinical significance of findings, the latter being the focus of the current study.

In 2021, a UK research group reported results from an uncontrolled study of psychological outcomes over 3 years (Carmichael et al., 2021a). However, unlike previous research groups, the data was made available through the UK Data Archive, reflecting current good practice in clinical outcomes research. This provides a unique opportunity to address the concern raised by the NICE review that studies of psychological outcomes in this field have omitted to assess clinically meaningful change.

The UK study received ethical approval from the UK National Research Ethics Service in 2011 and recruitment of participants took place from 2011 to 2014 (Carmichael et al., 2021a). The aims of the study were “to evaluate the benefits and risks for physical and mental health and wellbeing of mid-pubertal suppression in adolescents with GD; to add to the evidence base regarding the efficacy of GnRHa treatment for young people with GD; and to evaluate continuation and discontinuation of GD and the continued wish for gender reassignment within this group”. The study assessed physical response to pubertal suppression; bone health; adverse events; and psychological outcomes. Psychological outcomes included general psychopathology, self-harm, quality of life, body image, GD, general functioning and patient experience. General psychopathology was assessed using the CBCL (parent-report) and YSR (self-report). These are validated measures that have been widely used internationally to assess children’s mental health (Achenbach & Rescorla, 2001) and were used by two of the studies reviewed by NICE. These measures are arguably the most reliable indicator of general mental health and psychological functioning among the outcomes assessed in the study and because of available psychometric properties and normative data, are amenable to analysis of Reliable and Clinically Significant Change.

Anonymised data from the GIDS study were lodged at the UK Data Archive (Carmichael et al., 2021b) and can be downloaded for researcher use without any further additional ethical approval required. The aim of the current study was therefore to re-analyse the data from the GIDS study to assess Reliable and Clinically Significant Change on the CBCL and YSR for the sample at 12, 24 and 36 months follow-up.

## Materials and Methods

### Design

The current study is a secondary analysis of data collected at the UK GIDS service and stored at the UK Data Archive. The design of the current study is an assessment of individual level change using Reliable and Clinically Significant Change analysis (Jacobson & Truax, 1991). The original study was an uncontrolled pre-post design.

### Participants

In the original study, children meeting the eligibility criteria were referred by GIDS to University College London Hospital NHS Foundation Trust between 2011-2014, where study information was given and consent taken. Eligibility criteria were: age 12-15; seen by GIDS for at least 6 months; at least 4 assessment sessions in GIDS; psychologically stable; meeting criteria for GD (including high likelihood of extreme psychological distress with ongoing pubertal development); actively requesting GnRHA; capacity for informed consent; established puberty; and normal endocrine function. Forty-four children consented to take part in the study. Detailed study procedures and exclusion criteria are provided in the published paper (Carmichael et al., 2021a). Participants completed the YSR at baseline (n=44), 12 months (n=41) and 24 months (n=15). Parents completed the CBCL at baseline (n=43), 12 months (n=41), 24 months (n=20) and 36 months (n=11). All of the data available for these measures were included in the current analysis.

### Measures

The data deposited were standardised for age and sex and included individual level scores for CBCL Externalising, CBCL Internalising, CBCL Total Problems, YSR Externalising, YSR Internalising and YSR Total Problems. These sub-scales are “higher-order” subscales which combine other subscales into broad dimensions (Achenbach & Rescorla, 2001). The Internalising domain is considered a measure of general emotional problems including anxiety, depression, somatic complaints and being withdrawn. The Externalising domain is considered an aggregate scale of behavioural problems including attention problems and aggressive behaviour. The Total Problems score is considered an aggregate of emotional and behavioural problems as well as sleep difficulties (Guerrera et al., 2019). The scales have published psychometric properties including good internal reliability with Chronbach alphas ranging from 0.90 to 0.97 (Achenbach & Rescorla, 2001). The scales come from a broad system of measures (ASEBA) developed in the USA which have been widely used with published norms and reliability data and clinical cut-offs for all versions. Established cut-points are <60 (normal range), 60-63 (borderline range) and >63 (clinical range) (Achenbach & Rescorla, 2001).

### Reliable and Clinically Significant Change analysis

Comparing group averages using statistical tests is widely used in psychology experiments and outcome studies of clinical interventions. However, used alone in clinical research, the approach can be misleading. Finding a “statistically significant” difference between two average scores can lead people to believe that the difference (or lack of difference) is important in clinical terms (Field, 2013). Reporting effect sizes is one important additional statistic in clinical studies that helps determine the clinical significance of a result. However, like statistical significance of mean differences, the effect size still relies on group averages. Looking only at group averages and comparing them masks variation. Analysis of Reliable and Clinically Significant Change (Jacobsen & Truax, 1991) is an established complementary approach in clinical research which allows us to understand more about what is happening for individuals in a group rather than only what is happening “on average” for the whole group.

Analysing “reliable change” and “clinically significant change” (Blampied, 2022) uses the logic of individual change in which the number of people in the study does not influence whether the results are “statistically significant”. The approach relies on known psychometric features of the questionnaire being used, such as how reliable it is when used with similar groups of patients with similar problems and how it performs statistically in people with and without symptoms. Many people will have taken part in prior testing of the questionnaire to establish these features and properties, as is the case with the CBCL and YSR (Achenbach & Rescorla, 2001). Information about the questionnaire is used to provide parameters for analysis of individual change without requiring a minimum sample size for power or statistical significance.

The approach enables analysis of individual change within the group, rather than averaging out scores across the group which is a fairly narrow way to interpret results from clinical studies. Rather than saying “on average” there was a statistically significant difference before and after treatment; we can say “of the whole sample, 50% improved but 50% deteriorated” and so on. As such, evaluation of both adult and child psychological services in the UK use approaches which examine data at the level of individual change rather than group means (Gibbons, Harrison, & Stallard, 2021; Wolpert et al., 2016). These approaches typically assess two aspects of individual change: Reliable Change and Clinically Significant Change, the latter sometimes referred to as “recovery”.

Reliable Change is individual change that is sufficiently unlikely to have arisen by measurement error alone. The approach provides summary information about what proportion of the sample improve, deteriorate or stay the same (no change). The formula takes into account the standard error of difference (before and after treatment) as well as the internal reliability of the measure (Evans, Margison, & Barkham, 1998). The analysis is “applicable, in one form or another, to the measurement of change on any continuous scale for *any* clinical problem”(Evans et al., 1998). The approach can be used with clinical outcome data whether as part of a controlled or uncontrolled research study, or as part of routine outcome evaluation in a clinical setting. Clinically significant change or “recovery” refers to the proportion of patients who are within the clinical or borderline range at baseline, show reliable improvement and move into a non-clinical range (Gibbons et al., 2021; Jacobson & Truax, 1991).

The Reliable Change Index (RCI) for each sub-scale was calculated using the formula provided by Evans, Margison and Barkham using Chronbach alpha as the reliability criterion (Evans et al., 1998). Using the RCI, we calculated the proportion of participants in each of the three categories: No change, Deteriorate, Improve.

Clinically significant change (“recovery”) was calculated using the published clinical cut-points. Following Gibbons, Harrison and Stallard (2021), participants who scored in the borderline (subclinical) or clinical range at baseline (greater than or equal to 60) were included in the analysis of clinically significant change. Of those meeting this criterion, we calculated the proportion who had moved into the normal range at each time point.

### Case-by-case analysis

In order to examine patterns of change by individual, a case-by-case analysis was constructed in which each case is presented separately on one line (see Figure 1). The data were inspected to identify any observable patterns of individual change and cases were clustered according to patterns observed. This exercise did not involve any statistical testing given the small sample size; the method involved clustering cases based on qualitative observations only in order to inform further discussion.

**Figure 1.**
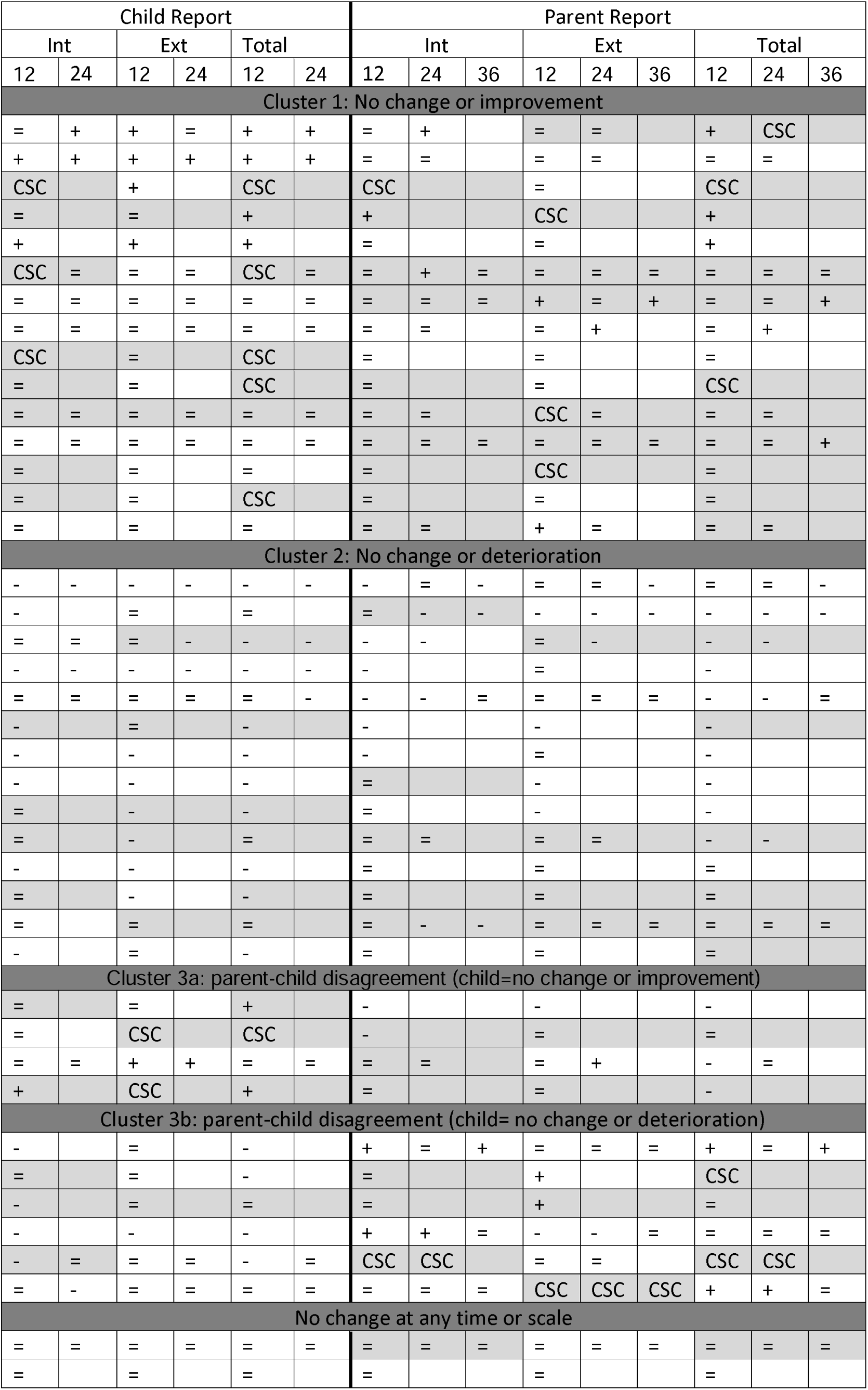
Case-by-case change patterns. notes: Int = Internalising Problems; Ext = Externalising Problems; Total = Total Problems; 12 = 12 months; 24 = 24 months; 36 = 36months; “=” = No Change; “+” = Reliable improvement; “-“ = Reliable deterioration; CSC = Reliable improvement and Clinically Significant Change. Shaded cells indicate the baseline score was in the borderline or clinical range Alt Text: a matrix with 41 rows and 15 columns in which each row is a participant and columns show parent-report and child-report by scale (Internalising Problems, Externalising Problems, Total Problems) and timepoint (12, 24 and 36 months). Cells contain plus, minus or equals symbols to show improvement, deterioration, or no change. Rows are divided into clusters: about a third of rows show plus and equals symbols only and about a third of rows show minus and equals symbols only. Remaining rows show different patterns according to child and parent report.

### Ethics

Data from the original study were anonymised and lodged at the UK Data Archive in a format suitable for use with no further ethical approval or consent required, as agreed with the Health Research Authority (Carmichael et al., 2021b). The original publication provides more detailed information on study methodology and further details on the ethics approval which was obtained from the National Research Ethics Service (NRES: reference 10/H0713/79) in February 2011 (Carmichael et al., 2021a). Subsequently, the team had discussions with the Health Research Authority who provided permission for data to be deposited with the UK Data Archive on the condition that sensitive data were removed to minimise disclosure risk of personal information (Carmichael et al., 2021a).

## Results

The findings for Reliable Change and Clinically Significant Change are presented below by Internalising Problems (Table 1), Externalising Problems (Table 2) and Total Problems (Table 3). [Insert Tables 1, 2 and 3]

**Table 1.** CBCL and YSR Internalising Scales – reliable change and reliable recovery.

| **<u>CBCL Internalising</u>** |  |  |  | **<u>YSR Internalising</u>** |  |  |
| --- | --- | --- | --- | --- | --- | --- |
| Reliable change | 12m | 24m | 36m | Reliable change | 12m | 24m |
| No change | 28 (68%) | 12 (60%) | 7 (64%) | No change | 23 (56%) | 10 (67%) |
| Deteriorate | 8 (20%) | 4 (20%) | 3 (27%) | Deteriorate | 12 (29%) | 3 (20%) |
| Improve | 5 (12%) | 4 (20%) | 1 (9%) | Improve | 6 (15%) | 2 (13%) |
| N | 41 | 20 | 11 | N | 41 | 15 |
| Clinical/borderline range at baseline | Reliable and Clinically Significant Change* |  |  | Clinical/borderline range at baseline | Reliable and Clinically Significant Change* |  |
| 24/43 | 2/22 | 1/11 | 0/6 | 20/44 | 3/17 | 0/3 |
| 56% | 9% | 9% | 0% | 45% | 18% | 0% |

\*Denominator is number of paired cases for the relevant time point which were in the borderline or clinical range at baseline

**Table 2.** CBCL and YSR Externalising Scales – reliable change and reliable recovery.

| **<u>CBCL Externalising</u>** |  |  |  | **<u>YSR Externalising</u>** |  |  |
| --- | --- | --- | --- | --- | --- | --- |
| Reliable change | 12m | 24m | 36m | Reliable change | 12m | 24m |
| No change | 27 (66%) | 14 (70%) | 7 (64%) | No change | 25 (61%) | 10 (67%) |
| Deteriorate | 6 (15%) | 3 (15%) | 2 (18%) | Deteriorate | 9 (22%) | 3 (20%) |
| Improve | 8 (20%) | 2 (15%) | 2 (18%) | Improve | 7 (17%) | 2 (13%) |
| N | 41 | 20 | 11 | N | 41 | 15 |
| Clinical/borderline range at baseline | Reliable and Clinically Significant Change* |  |  | Clinical/borderline range at baseline | Reliable and Clinically Significant Change* |  |
| 16/43 | 4/15 | 1/9 | 1/5 | 12/44 | 2/11 | 0/2 |
| 37% | 27% | 11% | 20% | 27% | 18% | 0% |

\*Denominator is number of paired cases for the relevant time point which were in the borderline or clinical range at baseline

**Table 3.** CBCL and YSR Total Scales.

| **<u>CBCL Total</u>** |  |  |  | **<u>YSR Total</u>** |  |  |
| --- | --- | --- | --- | --- | --- | --- |
| Reliable change | 12m | 24m | 36m | Reliable change | 12m | 24m |
| No change | 20 (49%) | 12 (60%) | 6 (55%) | No change | 15 (37%) | 9 (60%) |
| Deteriorate | 12 (29%) | 4 (20%) | 2 (18%) | Deteriorate | 14 (34%) | 4 (27%) |
| Improve | 9 (22%) | 4 (20%) | 3 (27%) | Improve | 12 (29%) | 2 (13%) |
| N | 41 | 20 | 11 | N | 41 | 15 |
| Clinical/borderline range at baseline | Reliable and Clinically Significant Change* |  |  | Clinical/borderline range at baseline | Reliable and Clinically Significant Change* |  |
| 25/43 | 4/23 | 2/11 | 0/5 | 19/44 | 6/17 | 0/3 |
| 58% | 17% | 18% | 0% | 43% | 35% | 0% |

\*Denominator is number of paired cases for the relevant time point which were in the borderline or clinical range at baseline

### Reliable Change

Data indicate that across all scales with both self-report (YSR) and parent report (CBCL), the majority of participants experience no reliable change in distress across all time points. Between 15% and 34% reliably deteriorate and between 9% and 29% reliably improve.

### Clinically Significant Change

As indicated in Tables 1, 2 and 3, there are relatively small numbers of participants in the clinical range at baseline for all subscales: 27% to 58% of the overall sample, depending on the subscale and parent versus child-report. Rates of reliable recovery range from 0% to 35% depending on the scale and time point. Reliable recovery fell to zero percent on all subscales at the final time point with the exception of CBCL Externalising scale which shows 1 out of 5 participants (20%) had moved into the normal range at 36 months.

### Case-by-case analysis

Figure 1 presents a case-by-case analysis in which each row represents one participant. Each row shows the direction of change for all scales and time points for both child and parent-report. Three main clusters of cases were identified based on observed patterns of change. Cluster 1 (n=15) represents cases in which individual level change fluctuated between reliable improvement and no change according to both parent-report and child-report. Cluster 2 (n=14) represents cases in which individual level change fluctuated between reliable deterioration and no change according to both parent and child report. Cluster 3 represents cases in which the pattern of change appears to be different depending on whether we look at parent-report or child-report. In cluster 3a (n=4), the child-report scales show fluctuation between no change and reliable improvement while the parent-report scales fluctuate mostly between no change and reliable deterioration. In cluster 3b (n=6), the child-report scales show fluctuation between no change and reliable deterioration while the parent-report scales mostly show fluctuation between no change and reliable improvement. Two cases do not fall into any of these clusters as they show no change across all time points and scales.

## Discussion

The original report of the GIDS study found no statistical differences on the CBCL or YSR between time points, concluding that either puberty blockers “brought no measurable benefit nor harm to psychological function”; or that “treatment reduced [the] normative worsening of problems”. The latter conclusion is based on evidence that psychological problems as measured by the YSR and CBCL tend to worsen during early adolescence, which is one way to interpret the findings. However, the evidence cited comes from non-clinical populations (Verhulst, 2003). Children who attend GIDS are a clinical population (Di Ceglie et al., 2002; Holt et al., 2016), with a similar range of social and psychological problems to those seen in CAMHS populations. Moreover, the children in the GIDS study were receiving mental health support from the GIDS service in a therapeutic setting, making them a clinical population by definition. Therefore, an alternative benchmark applied in our discussion of findings below is the UK CAMHS population rather than the general population (non-clinical). When evaluating CAMHS services in the UK, it is standard practice to examine individual level change using Reliable and Clinically Significant Change methods and to benchmark interpretations against CAMHS psychological outcomes.

The analysis is limited by absence of UK norms for the measures used, combined with the lack of a study control group to provide local norms. Clinical cut points used were based on US normative data and it is possible that UK teenagers may present differently to US teenagers in respect of GD. The ASEBA tools have no specific normative data for children with GD, although as noted earlier, UK GD clinic populations present with psychological profiles broadly similar to those of UK CAMHS populations. It would therefore be useful for any future clinical studies to use outcome measures commonly used for outcome monitoring and reliable change analysis in UK CAMHS populations such as the RCADS (Goodman, 1997) or SDQ (Chorpita, Yim, Moffitt, Umemoto, & Francis, 2000). The analysis is limited by the limitations of the dataset available for re-analysis which lacks differentiation by sex and item level data to look at more fine-grained sub-scales. The original study was uncontrolled and has a small sample size. The very small sample size at 36 months is particularly important to bear in mind when interpreting results. The reason for the reduction in sample size is given as “people were recruited at different ages (12–15 years) but left the study soon after their 16^th^ birthday”; no further explanation is given about why this was the case (Carmichael et al., 2021a). While the individual level change approach can be applied irrespective of sample size and lack of a controlled design, there remains a fundamental problem that there is insufficient data to draw firm conclusions about the safety and efficacy of this treatment. It is therefore important that any future studies should be sufficiently powered and designed to minimise data loss when the treatment stops, either because of moving to a different treatment or stopping treatment.

Nevertheless, this is the first analysis of data on children aged 12-15 with GD taking puberty blockers demonstrating individual level change as opposed to testing differences between group averages. As such this analysis gives a fuller picture of the benefits and risks of treatment. This secondary analysis of Reliable and Clinically Significant Change of the UK GIDS data indicates that, broadly speaking, for Internalising and Externalising Problems, 37%-70% experience no reliable change in distress across time points. Between 15% and 34% deteriorate; and between 9% and 29% reliably improve. The Total Problems scale shows higher proportions deteriorating (18%-34% depending on time point).

Observed rates of reliable improvement are not dissimilar to other CAMHS service evaluations. For example, using a similar global scale of psychopathology (RCADS), Gibbons, Harrison and Stallard (Gibbons et al., 2021) found between 20.7% and 24.4% of their sample reliably improved. However, the same study found that nearly all the remaining participants showed No Change with only a very small proportion (0.7% - 5.7%) deteriorating. These proportions were similar for a range of other psychological outcome variables used (Gibbons et al., 2021). Comparatively high levels of deterioration in the GIDS sample (ranging from 15-34%) is therefore concerning. It is important to note that the highest rate of deterioration (34%) is seen in the self-report scale at 12 months where the largest sample size is available.

Case-by-case analysis of patterns of change across individuals indicates that about a third of participants fluctuate between no change and improvement; and about a third fluctuate between no change and deterioration according to both parent-report and child-report. For about a quarter of participants (ten), the pattern of change reflects disagreement between parent-report and child-report. This may point to the importance of clinical work to identify families where there may be different understandings of the child’s difficulties and provide more in-depth family work in these cases. Relatively few participants fell into the borderline or clinical range at baseline (27%-58% depending on the scale). This is lower than a typical CAMHS population; Gibbons, Harrison and Stallard (2021) found around 90% of their sample to be in the clinical or borderline range at baseline. This suggests that the GIDS participants may have been less distressed when they were referred to the clinic than typical CAMHS referrals and may indicate a relatively low referral threshold operating at the time period studied.

Given the relatively low proportion of the sample in the clinical or borderline range, the rates of clinically significant change should be treated with caution. Nevertheless, a notable finding is that the proportion showing reliable and clinically significant change reduces over time so that at the later measurement points, self-report and parent-report rates of clinically significant change converge at zero, with the exception of parent-report externalising problems which shows 20% (1/5) moved to the normal range at 36 months. CAMHS services typically see rates of clinically significant change around 50% (Gibbons et al., 2021) and therefore the rates seen in the GIDS sample are comparatively low.

Using the reliable and clinically significant change approach to analysis of clinical study data provides an opportunity for research teams in this field to conduct fuller analysis of their data to ascertain whether there are any variables which might predict which children with GD are most likely to benefit psychologically and which are most likely to deteriorate, rather than considering the group as uniform in likely response to treatment. Statistical significance testing relies on minimum sample sizes which have been difficult to obtain in this field to date. This complementary analytic approach allows us to look at how a treatment is performing in terms of the percentage of patients improving, deteriorating and showing clinically significant change. Larger samples are absolutely necessary irrespective of whether group level analysis or individual level analysis (RCSC) is applied. However, it is possible, using this approach, to look at patterns, such as who is benefitting and who is not to inform practice and research in the interim. For example, the approach could be used to look more closely at biological sex in order to say, for example, that “x% of biological boys improved”; “x% of biological girls deteriorated”; “x% of biological boys experienced side effect A” and so on, which would improve consent processes in clinical practice until better data is available. Many studies in this field have small sample sizes, limiting statistical testing within groups by sex. In the absence of adequate data, coupled with children accessing the treatment through private prescriptions, using analysis of individual change to observe patterns like this would be useful to guide further research, help inform parents and children more clearly, and lead to more individualised, personalised care. The approach could also enable comparisons in the absence of formal control groups. For example, it would be possible to analyse routinely collected outcome data for children who do not take up puberty blockers to generate comparable data on Reliable and Clinically Significant Change as a naturalistic control group. We recommend that these approaches be incorporated into new GD services being established in the UK as well as new research studies being designed.

## Data Availability

The data underlying the results presented in this study are available from the UK Data Service (DOI: 10.5255/UKDA-SN-854413).

https://doi.org/10.5255/UKDA-SN-854413

## References

Achenbach, T. M., & Rescorla, L. A. (2001). Manual for the ASEBA School-Age Forms & Profiles. Burlington, VT: University of Vermont, Research Center for Children, Youth, & Families.

Barnes, H. (2023). Time to Think: The Inside Story of the Collapse of the Tavistock’s Gender Service for Children. London: Swift Press.

Blampied, N. M. (2022). Reliable change and the reliable change index: still useful after all these years? The Cognitive Behaviour Therapist, 15, e50. 10.1017/S1754470X22000484

Carmichael, P., Butler, G., Masic, U., Cole, T. J., De Stavola, B. L., Davidson, S., … Viner, R. M. (2021a). Short-term outcomes of pubertal suppression in a selected cohort of 12 to 15 year old young people with persistent gender dysphoria in the UK. PLOS ONE, 16(2), e0243894. 10.1371/journal.pone.0243894

Carmichael, P., Butler, G., Masic, U., Cole, T. J., De Stavola, B. L., Davidson, S., … Viner, R. M. (2021b). Short-term outcomes of pubertal suppression in a selected cohort of 12 to 15 year old young people with persistent gender dysphoria in the UK 2019-2021 [Data Collection]. 10.5255/UKDA-SN-854413

Chorpita, B. F., Yim, L., Moffitt, C., Umemoto, L. A., & Francis, S. E. (2000). Assessment of symptoms of DSM-IV anxiety and depression in children: A revised child anxiety and depression scale. Behaviour Research and Therapy, 38(8). 10.1016/S0005-7967(99)00130-8

Cohen-Kettenis, P. T., & van Goozen, S. H. M. (1998). Pubertal delay as an aid in diagnosis and treatment of a transsexual adolescent. European Child & Adolescent Psychiatry, 7(4), 246–248. 10.1007/s007870050073

Costa, R., Dunsford, M., Skagerberg, E., Holt, V., Carmichael, P., & Colizzi, M. (2015). Psychological Support, Puberty Suppression, and Psychosocial Functioning in Adolescents with Gender Dysphoria. The Journal of Sexual Medicine, 12(11), 2206–2214. 10.1111/jsm.13034

de Vries, A. L. C., Steensma, T. D., Doreleijers, T. A. H., & Cohen-Kettenis, P. T. (2011). Puberty Suppression in Adolescents With Gender Identity Disorder: A Prospective Follow-Up Study. The Journal of Sexual Medicine, 8(8), 2276–2283. 10.1111/j.1743-6109.2010.01943.x

Di Ceglie, D., Freedman, D., McPherson, S., & Richardson, P. (2002). Children and adolescents referred to a specialist gender identity development service: Clinical features and demographic characteristics. International Journal of Transgenderism, 6(1).

Evans, C., Margison, F., & Barkham, M. (1998). The contribution of reliable and clinically significant change methods to evidence-based mental health. Evidence-Based Mental Health, 1(3), 70–72. 10.1136/ebmh.1.3.70

Field, A. (2013). Discovering Statistics Using IBM SPSS Statistics (4th ed.). Sage Publications Ltd.

Gender Identity Development Service. (n.d.). The Early Intervention Study. Retrieved from https://gids.nhs.uk/research/early-intervention-study

Gibbons, N., Harrison, E., & Stallard, P. (2021). Assessing recovery in treatment as usual provided by community child and adolescent mental health services. BJPsych Open, 7(3), e87. 10.1192/bjo.2021.44

Goodman, R. (1997). The strengths and difficulties questionnaire: A research note. Journal of Child Psychology and Psychiatry and Allied Disciplines, 38(5). 10.1111/j.1469-7610.1997.tb01545.x

Guerrera, S., Menghini, D., Napoli, E., Di Vara, S., Valeri, G., & Vicari, S. (2019). Assessment of Psychopathological Comorbidities in Children and Adolescents With Autism Spectrum Disorder Using the Child Behavior Checklist. Frontiers in Psychiatry, 10. 10.3389/fpsyt.2019.00535

Holt, V., Skagerberg, E., & Dunsford, M. (2016). Young people with features of gender dysphoria: Demographics and associated difficulties. Clinical Child Psychology and Psychiatry, 21(1), 108–118. 10.1177/1359104514558431

Jacobson, N. S., & Truax, P. (1991). Clinical significance: A statistical approach to defining meaningful change in psychotherapy research. Journal of Consulting and Clinical Psychology, 59(1), 12–19. 10.1037/0022-006X.59.1.12

National Institute for Health and Care Excellence. (2020). Evidence review: Gonadotrophin releasing hormone analogues for children and adolescents with gender dysphoria. Retrieved from https://www.engage.england.nhs.uk/consultation/puberty-suppressing-hormones/user_uploads/nice-evidence-review-gnrh-analogues-for-children-and-adolescents-with-gender-dysphoria-october-2020.pdf

NHS England. (2023). URN 1927 Literature Surveillance Report. Retrieved from https://www.engage.england.nhs.uk/consultation/puberty-suppressing-hormones/user_uploads/literature-surveillace-report-on-gnrh-analogues-for-children-and-adolescents-with-gender-dysphoria-may-2023.pdf

Staphorsius, A. S., Kreukels, B. P. C., Cohen-Kettenis, P. T., Veltman, D. J., Burke, S. M., Schagen, S. E. E., … Bakker, J. (2015). Puberty suppression and executive functioning: An fMRI-study in adolescents with gender dysphoria. Psychoneuroendocrinology, 56, 190– 199. 10.1016/j.psyneuen.2015.03.007

Wolpert, M., Jacob, J., Napoleone, E., Whale, A., Calderon, A., & Edbrooke-Childs, J. (2016). Child- and Parent-Reported Outcomes and Experience from Child and Young People’s Mental Health Services 2011–2015. Retrieved from https://www.corc.uk.net/media/1544/0505207_corc-report_for-web.pdf

